# Sheltering in place and the likelihood of non-natural death

**DOI:** 10.1101/2020.12.15.20248261

**Authors:** Ralph Catalano, M. Maria Glymour, Yea-Hung Chen, Kirsten Bibbins-Domingo

## Abstract

Increasing hospitalizations for COVID-19 in the United States (US) and elsewhere have ignited debate over whether to reinstate shelter-in-place policies adopted early in the pandemic to slow the spread of infection. The debate includes claims that sheltering in place influences deaths unrelated to infection or other natural causes. Testing this claim should improve the benefit/cost accounting that presumably informs the decision of whether to reimpose sheltering in place. To distinguish effects of shelter-in-place policies from other events in the pandemic, we compare experiences in two large US states with markedly different policies. We use time-series methods to compare temporal variation in non-natural deaths in California to that in Florida. California was the first state to begin and among the last to end sheltering in place while sheltering began later and ended earlier in Florida. We find that during weeks when California had shelter-in-place orders in effect, but Florida did not, the odds that a non-natural death occurred in California rather than Florida fell 14.8% below values expected from history. These results suggest that sheltering-in-place policies reduce mortality from mechanisms unrelated to infection or other natural causes of death.

**Significance Statement:** We address what has become an unusually important, but contentious, question – Did mandated sheltering in place affect the incidence of non-natural death in the United States? We use time-series methods to exploit a “natural experiment” in which California and Florida imposed and relaxed stay-at-home orders at different times. We find that shelter-in-place orders likely reduced non-natural mortality. We argue that ignoring averted non-natural deaths will lead not only to an underestimate of the benefits of the shelter-in-place policies, but also to an undercount of the impact of COVID-19 infections on natural deaths.

---

As COVID-19 infections and hospitalizations increase in the US and elsewhere, debate over the prudence of shelter-in-place mandates has intensified (1, 2). The debate includes claims that the intervention has induced unintended adverse health effects (3). The “Great Barrington Declaration,” for example, justifies opposition to sheltering in place by citing “devastating effects on short and long-term public health (2, 4).” Others have extended this argument to suggest that harms of lockdowns include increases in deaths not just from impeded access to health care, but also from suicide (5) and intrafamily violence (6).

Although clinical anecdote (7) supports the intuition of increased mortality during sheltering-in-place, the scholarly literature reports mixed results (8–10). These divergent results likely arise, at least in part, because the observed populations varied in the fraction at risk of what the Centers for Disease Control (CDC) term “natural” death or that plausibly averted had sheltering in place not disrupted medical care (11). Although information characterizing deaths varies among states, all death certificates in the US use a “manner of death” classification that includes “natural death” defined as ‘‘due solely or nearly totally to disease and/or the aging process (11).’’ Nearly all deaths attributed to Covid-19, for example, should appear in the “natural death” category. Based on data from the last decade, natural deaths account for approximately 89% of deaths in the US (12).

The observed populations also likely varied in the fraction at risk of “non-natural” deaths. How sheltering in place affects these deaths remains, however, even less clear than its effect on natural deaths. As noted above, anecdote implies that suicides (5) and death due to intrafamily violence (7) increased with sheltering in place. Other literature, however, reports decreases in such non-natural deaths as those by accidents (13), stranger-on-stranger violence (14), and medical error (15). Taken together, these reports raise an important, question -- what has been the “net effect” of sheltering in place on non-natural deaths? Answering this question requires an estimate of the association between sheltering in place and the incidence of non-natural deaths. The peer-reviewed literature, however, includes no attempts to estimate that association. We attempt such an estimate using data from two US states. We use time-series methods to compare non-natural deaths in California, the first state to begin and among the last to end sheltering in place, with those in another large state, Florida, where sheltering in place began late and ended early.

California and Florida responded very differently to the emerging epidemic in early 2020. Large employers, primarily in the technology sector, began telling their California workers to stay at home effective March 8 (16). Counties in the San Francisco Bay Area issued stay-at-home orders effective March 17 (17) and the Governor issued similar orders for the remainder of the state on March 19 (18). California began reopening on May 15 (19). Florida mandated sheltering in place effective April 3 and reopened on May 1 (20, 21). Data that indirectly measure the behavior of households suggest that the citizens of both states reduced mobility and social contacts when shelter-in-place orders were in effect and increased both when orders were removed (22).

## METHODS

### Data

CDC regularly publishes weekly death counts by state and select causes from 2014 through the most recently accounted week (23). Our tests used deaths in California and Florida for 333 weeks beginning December 29, 2013 (first data available) and ending May 16, 2020 (last week of sheltering in place in California). We subtracted weekly “Natural Cause” deaths from total deaths to estimate the incidence of non-natural deaths in both states. These deaths include, as suggested above, death by accident, suicide, interpersonal violence, drug overdoses, and medical error (i.e., iatrogenesis). State counts of 2020 non-natural deaths by subcategories have not yet been published. In 2019, however, motor vehicle crashes and suicide each accounted for approximately 16% of non-natural deaths in both California and Florida while homicide contributed about 6.5% in each state (12).

We computed the weekly odds that a non-natural death in either state occurred in California. We transformed those odds to natural logarithms (i.e., logits) to allow us to express results as percent difference between observed and expected odds during weeks when California alone sheltered in place.

### Analyses

The weekly odds that a death occurred in California instead of Florida exhibit patterns over time (i.e., autocorrelation) that can lead to an expected or predicted value different from the mean of all weeks. As described below, we identify and model autocorrelation in the logits before shelter in place orders and develop predictions for ensuing weeks. We then ask whether the observed values differed from predicted during periods when shelter-in-place orders were in place in California but not Florida.

Our test proceeded through 5 steps consistent with time-series conventions used in epidemiology (24,25). First, we used Box-Jenkins methods (26) to identify and model autocorrelation in a 323-week “training period” up to March 8, 2020. Second, we used the training model to forecast values for the next 10 weeks. Third, we created a 333 week “counterfactual series” by joining the 10 forecasts from step 2 to the 323 fitted values of the training model estimated in step 1. Fourth, we subtracted the 333 counterfactual logits from the observed logits to estimate a residual series. The residual series measure the degree to which the likelihood of a non-natural death in California differed from that expected based on such deaths in Florida and on historical patterns (i.e., autocorrelation). Fifth, we regressed the residual series computed in Step 4 on a binary shelter-in-place variable scored 1 for the weeks when California had shelter-in-place orders in effect but Florida did not, and scored 0 for all other weeks. The variable, therefore, equaled 1 for the 3 weeks starting March 8 and ending March 28 and for the 3 weeks starting May 3 and ending May 16.

If sheltering in place changed the likelihood of non-natural deaths, the coefficient for the shelter-in-place variable would appear detectably different from 0. We set detection sensitivity at P< .05. Subtracting the antilog of that coefficient from 1 and multiplying the difference by 100 yields the percent difference between observed and expected odds.

## RESULTS

Weekly incidence of non-natural deaths ranged from 210 to 511 (mean 399) in California during our test period, and from 176 to 452 (mean 340) in Florida. Figure 1 shows the deaths plotted by week.

**Figure 1.**
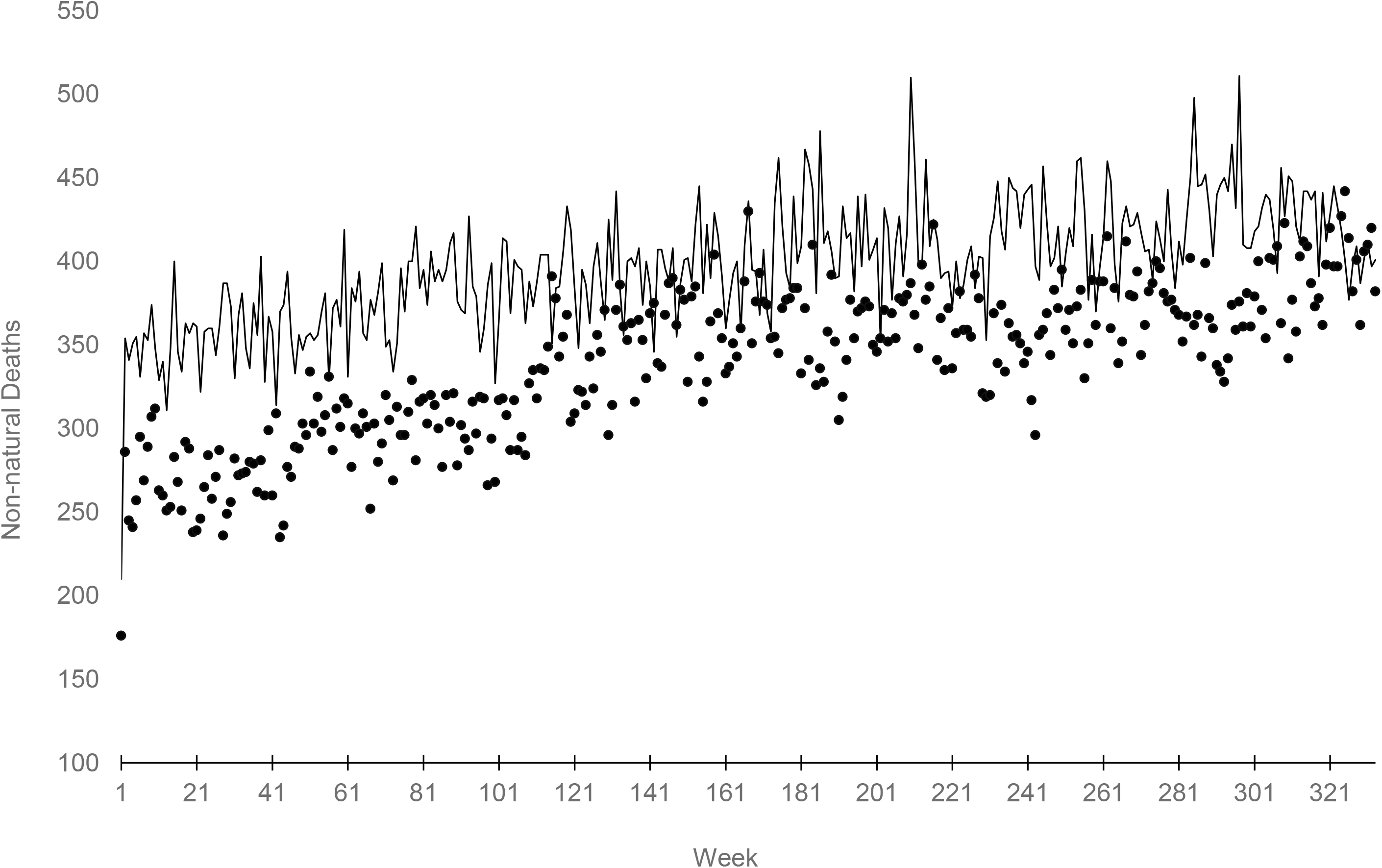
Weekly counts of non-natural deaths in California (line) and Florida (points) for 333 weeks starting December 29, 2013 and ending May 16, 2020.

The best fitting Box-Jenkins model, identified and estimated in Step 1, for logits in the 323-week training period was as follows:

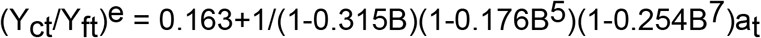

(Y_ct_/Y_ft_)^e^ is the log odds that a non-natural death in California or Florida during week t occurred in California. The constant, 1.184 (SE 0.016), equals the mean of (Y_ct_/Y_ft_)^e^. The 3 autoregressive parameters (i.e., 0.315, 0.176, and 0.254), all of which exceeded at least twice their standard errors (i.e., 0.054, 0.057, and 0.056 respectively), imply that including the values of (Y_ct_/Y_ft_)^e^ at weeks t-1, t-5 and t-7 improves, over using only the mean of past values, the model’s prediction of (Y_ct_/Y_ft_)^e^ at week t. B is the “backshift operator” or value of (Y_ct_/Y_ft_)^e^ at week t-1, t-5, or t-7. a_t_ is the model error term at week t.

The results of Steps 2 and 3, in which we construct a counterfactual series by joining values forecasted for weeks 324 through 333 to the fitted values during to the training weeks, appear in Figure 2 as a line. We show only the last year of data to allow better resolution of the information. The points in Figure 2 show the observed values for the 333 weeks. The residual series, computed in Step 4 by subtracting the full 333 expected from observed values, became the dependent variable for our test.

**Figure 2.**
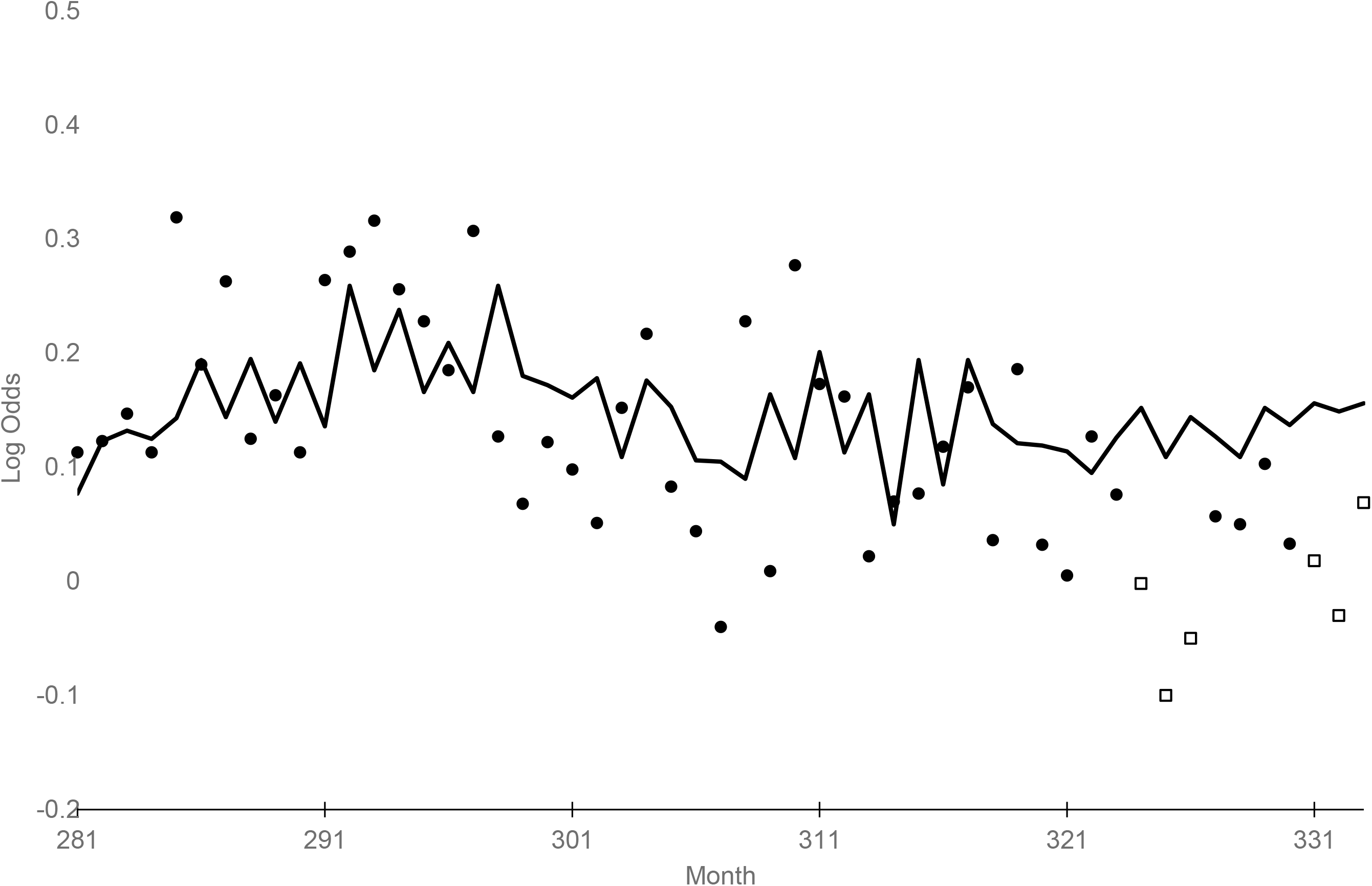
Expected (line) and observed (points) weekly log odds that a non-natural death in California and Florida occurred in California. Boxes show weeks during which California but not Florida sheltered in place (n = 52 weeks beginning May 19, 2019 and ending May 16 2020).

Step 5, in which we regress the residual series on our shelter-in-place binary variable yielded a regression coefficient of -0.160 (SE 0.037). This estimate implies that the odds of a nonnatural death in the two states occurring in California detectably *decreased* by 14.8% when California alone sheltered in place.

The association between the start of sheltering in place and non-natural deaths may not persist throughout its imposition. To shed light on the week-by-week association, we repeated our main test but substituted a binary variable scored 1 for the week ending March 14 and 0 otherwise for the shelter in place variable described above. We estimated its association with our dependent variable in the synchronous configuration (i.e., both occurring in the same week) as well as in 9 lagged configurations in which the dependent variable ranged from 1 through 9 weeks after that ending March 14. Table 1 shows the results. The pattern of coefficients appears consistent with our main results in that greater differences between expected and observed appear when California alone sheltered in place. Stronger associations, however, appear in the weeks when California started sheltering in place and Florida had yet to start. The associations appear smaller when California remained sheltering in place and Florida reopened.

**Table 1.**
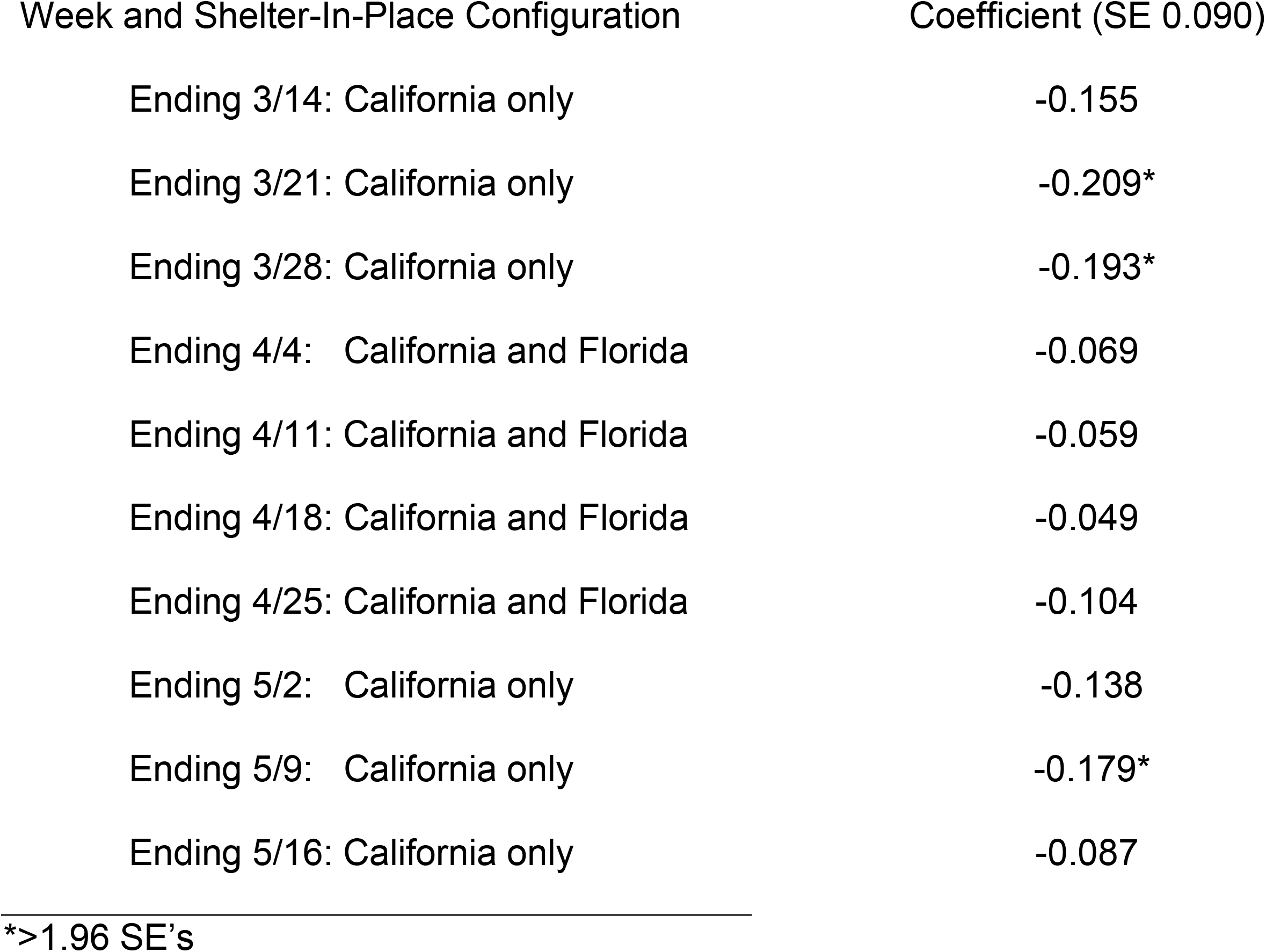
Coefficients from regression predicting observed less predicted (from autocorrelation) weekly log odds that a non-natural death in California and Florida occurred in California (n = 333 weeks beginning 12/29/2013 and ending 5/16/2020).

We tested whether our main finding would appear if we allowed the Box-Jenkins modeling in Step 1 to include all 333 months. This approach is “naïve” to the timing of the assumed interruption and therefore allows values during shelter in place to influence estimation of the Box-Jenkins parameters as well as subsequent counterfactuals. This test converged with our main test by detecting the same structure of autocorrelation and essentially the same association between the shelter-in-place variable and the odds of non-natural death occurring in California.

## DISCUSSION

Time-series modeling using seven years of weekly non-natural deaths shows that when California ordered sheltering in place but Florida did not, Californians yielded unexpectedly few non-natural deaths. We estimate an approximate 15% reduction below values expected from non-natural deaths in Florida and from historical patterns. The estimated benefit appears larger at the outset of mandated sheltering in place than near its suspension.

Our findings have implications for basic as well as and policy research. First, they imply that basic epidemiologic research intended to estimate deaths arising from impeded access, regardless of its source, to routine medical care should exclude non-natural deaths from tests of association. Including them may lead to underestimating the pathogenic effect of such impedance because medical care likely has relatively little effect on their incidence and because they may account for 10% or more of all deaths. This circumstance similarly implies that benefit/cost analyses intended to inform the policy debate over whether and when to employ sheltering in place to manage epidemics needs to use results from prior epidemiologic research with care. The scholarly work intended to estimate deaths accruing to impeded access to routine medical care has included non-natural deaths in the accounting. Using results from research that does not exclude those deaths could lead to an underestimate of deaths and their costs.

Advantages of our approach include that Box-Jenkins modeling accounts for autocorrelation including trends, cycles (e.g., seasonality), and the tendency for a series to remain elevated or depressed after high of low values. Using Florida as a comparison population, moreover, allows us to control for events, unrelated to shelter-in-place orders, that influenced non-natural deaths in both California and Florida.

Limitations of our approach include that currently available data do not allow us to distinguish among types of non-natural deaths. Further research should, when CDC publishes more detailed data, test the intuitive hypothesis that some types of these deaths, those by auto crash and medical error, for example, decrease during shelter in place while others, those by suicide and intrafamily violence for instance, may increase.

We do not claim a full accounting of the costs associated with non-natural deaths in California or Florida. We did not include, for example, the morbidity of non-fatal injuries averted by sheltering in place. CDC data indicate that for every 1 motor vehicle crash fatality, for example, there are 9 non-fatal hospitalizations and 88 individuals treated and released from emergency departments (27). These facts suggest that the reductions in non-natural fatalities that we measured trace much larger reduction in non-fatal morbidity and disability.

We did not attempt to estimate the association between non-natural deaths and social distancing induced by circumstances other than shelter-in-place orders. Mobility measured by household surveys or cellphone movement (22), for example, may vary directly with non-natural deaths. Future work should include such indicators of mobility to estimate the separate effects of differing interventions.

Evaluating the association between mandated sheltering-in-place policies and all deaths or non-Covid-19 deaths may provide a misleading picture of the benefits of these interventions. Shelter-in-place orders widely adopted in response to the COVID-19 epidemic likely reduced not only contagious illness but also non-natural mortality. Ignoring averted non-natural deaths will lead to an underestimate of the benefits of the shelter-in-place policies. Similarly, ignoring reductions in non-natural deaths may also lead to an undercount of the impact of COVID infections on death.

## Data Availability

All data used in these analyses can be accessed at publicly available websites cited in the manuscript.

## Notes

**Competing Interest Statement:** The authors declare no competing or financial interests.

### Competing Interest Statement

The authors have declared no competing interest.

### Clinical Protocols

https://data.cdc.gov/NCHS/Weekly-Counts-of-Deaths-by-State-and-Select-Causes/muzy-jte6)

### Funding Statement

No Funding

### Author Declarations

IRB exempt; uses only state-level vital statistics

